# Site-specific analysis of thoracic aortic aneurysm and cardiovascular mortality: Insights from the National Echo Database Australia

**DOI:** 10.1101/2025.04.22.25326179

**Authors:** James Nadel, Avan Suinesiaputra, Elizabeth D. Paratz, Juile Humphries, Alistair Young, Rene Botnar, David S. Celermajer, Geoff Strange, David Playford

**Author notes:** **Corresponding author:** Dr James Nadel, Heart Research Institute, 7 Eliza St. Newtown, New South Wales, Sydney, Australia, 2042. Twitter: @docnadel.

## Abstract

**Background:** Aortic diameter remains the most utilised criterion for considering surgical correction. In uncomplicated cases guidelines do not differentiate between the size of aneurysms at the root and ascending aorta. In order to improve practice, greater understanding of site-specific TAA is needed. A nationwide echocardiographic dataset linked to mortality outcomes was examined to determine how TAA affects cardiovascular (CVD) mortality.

**Methods:** The National Echo Database Australia (NEDA) was examined for aortic dimensions at the sinuses of Valsalva (SoV), sinotubular junction (STJ) and ascending aorta (AscAo). Patients were stratified according to absolute aortic diameters and grouped as normal (<4cm), mild (4.0-4.5cm), moderate (4.5-5cm) and severely (>5cm) dilated at the prescribed thoracic aortic sites. Mortality data was linked from the National Death Index.

**Results:** 477,501 echocardiographs from 175,158 patients with 2,897,357 patient-years of follow-up were included. Severe TAA at any site increased likelihood of 10-year CVD mortality compared to normal aortic diameters (31% vs. 14%, p<0.0001), with incremental increase in probability of CVD death when moving from the proximal to distal ascending aorta; CVD mortality at SoV 30% (HR 1.79; CI 1.2-2.67; p=0.004), STJ 41% (HR 1.91; CI 1.11-3.29; p=0.002) and AscAo 45% (HR 3.96; CI 2.06-7.64; p<0.001).

**Conclusions:** Severe TAA increases the probability of cardiovascular mortality. Given the low event rate of aortic death (0.2%) this is not solely explained by increased dissection risk. Interestingly, there is a doubling of CVD mortality likelihood when moving from the proximal to distal ascending aorta. These results suggest those with severe AscAo dilatation may be at higher CVD risk compared to those with aortic root aneurysms, identifying new considerations for risk stratification and surgical management.

## Introduction

Thoracic aortic aneurysms (TAA) manifest clinically as acute aortic syndromes with aortic dissection (AoD) comprising the majority of events.(1) AoD carries a high probability of fatality, reportedly up to 80% in sufferers.(2–4) Despite diagnostic advances, detection of TAA is challenging due in part to patients being asymptomatic prior to acute events and limitations in risk prediction to guide preventative surgical correction.(5–8) Owing to its high mortality and disability-adjusted life years, coupled with the need for routine multi-modal imaging surveillance and surgical correction, TAA and its sequelae represent a significant and underappreciated burden of disease.

The prevalence of AoD is increasing independent of an aging population,(9, 10) with incidence of acute thoracic AoD reported to range from 2.5 to 9 cases per 100,000 person-years.(11–13) Nevertheless, AoD is an under-recognised primary cause of death and thought to be routinely misclassified.(13, 14) The main treatment for TAA is prophylactic surgical repair and the decision to intervene is adjudged primarily on aortic size.(14–17) Acute complications are reported to be rare in TAA of moderate dilatation with a yearly dissection risk of 0.1%, 0.2%, and 0.6% at diameters of 4.5, 5, and 5.5 cm respectively, but incidence sharply rise to 7% per year when the diameter exceeds 6 cm.(18) In order to avoid aneurysmal expansion beyond these critical points, guidelines recommend preventative surgery at any site where the proximal ascending thoracic aortic diameter is >5-5.5 cm or lower in the presence of connective tissue disorders, a family history of AoD or concomitant bicuspid aortic valve morphology.(14–17) Interestingly, these guidelines do not differentiate between their recommendations at various sites along the proximal ascending aorta.

Despite a link between the severity of TAA and unfavourable outcomes, most dissections occur at diameters below guideline recommendations for prophylactic surgical correction. The International Registry of Acute Aortic Dissection (IRAD) a multinational database of over 7,300 aortic dissection cases found that 60% of patients had aortic diameters below the threshold for prophylactic surgery, suggesting that current guidelines designed to mitigate dissection fail to prevent the majority of events.(19) Additionally, since the aorta dilates directly following the onset of dissection,(20) it is likely that the number of events that would have been prevented based on current diameter cut-offs is even lower than those represented in IRAD. We have recently provided evidence to support this hypothesis in a large echocardiographic population, demonstrating that despite severe aortic aneurysms conferring a near 30-fold increase risk of aortic death, over 90% of the population who experienced mortality relating to aortic disease did so at diameters <5.5 cm prior to their event.(21)

This pattern has been described as ‘the aortic paradox’ and reinforces the core statistical principle that event rate is a function of both the likelihood of an event and the population numbers in which it occurs.(22) That is, despite a grossly increased probability of aortic dissection in severely dilated aortas, given their low prevalence, the much larger population who have non-severely dilated aortas experience the majority of fatal AoD. Nevertheless, it is unlikely that lowering the thresholds for surgical intervention alone would bear a long-term mortality benefit as it may expose a considerable number of patients with smaller aneurysms and thus, minimal yearly risk of dissection, to unnecessary elective surgery which in itself carries morbidity and mortality risk. As a result, research and clinical interest must shift to improving risk stratification metrics and enhancing criteria by which patients are selected for prophylactic surgical correction.

Using a nationwide echocardiographic (ECHO) database in a system of universal healthcare linked to mortality data, this study sought to examine how differing sites of TAA affects mortality over time with the aim of enhancing the prognostic understanding and management.

## Methods

### NEDA & the NDI

A multicentred retrospective cohort study was performed using the National Echo Database of Australia (NEDA). The protocols of NEDA have been described in detail elsewhere.(23–25) In short, NEDA is a multicentre observational database that captures reporting data from adults who have been referred for ECHO to >25 participating sites nationwide in a system of universal healthcare. The database includes national representation, is highly multicultural and sex-equitable with a mix of inpatient and outpatient echocardiograms for >600,000 adults and over 1 million ECHOs. Data from NEDA is routinely linked with the National Death Index (NDI) of Australia to obtain survival status, identifying fact, date and cause of death.

### Inclusion & exclusion criteria

At the time of study 1,048,575 ECHOs from 616,284 individuals were available in NEDA with the last NDI census date performed in May 2019. We included any patient with >1 ECHO, and following exclusion of duplicate studies, removal of data errors (any aortic measure <2 cm or >9 cm) and exclusion of studies where no aortic measurements were recorded, a cohort of 175,158 patients comprising 477,501 individual ECHOs and a total of 2,897,357 patient-years of follow-up were included for the final analysis (**Figure 1**).

**Figure 1:**
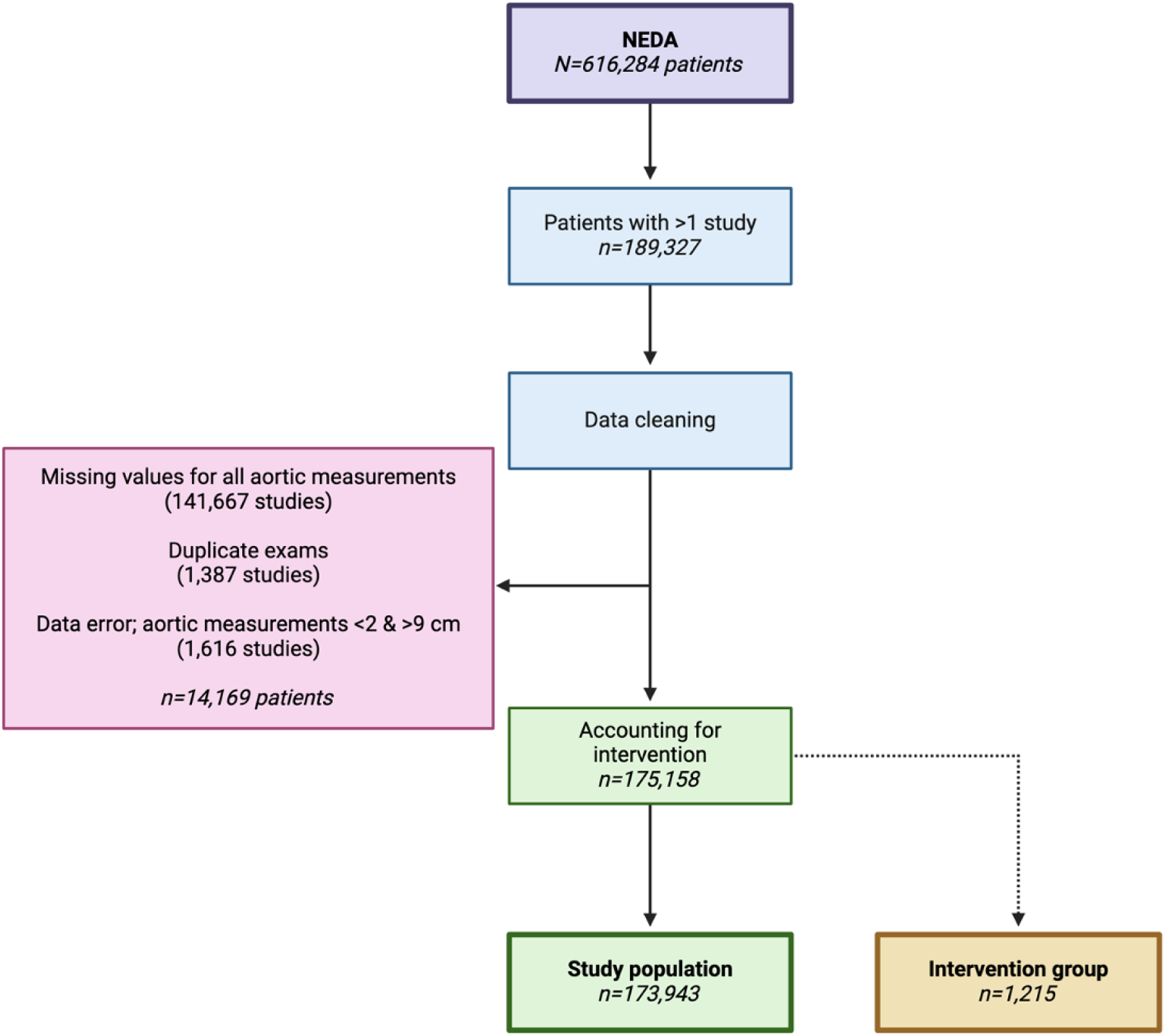
Inclusion criteria & data cleaning flowchart

### Defining aortic dimensions & groups

Echocardiographic measurements taken at sinuses of Valsalva (SoV), sinotubular junction (STJ) and ascending aorta (AscAo) were obtained and patients were grouped as having normal (<4 cm), mild (4-4.5 cm), moderate (4.5-5 cm) and severely (>5 cm) dilated thoracic aortas at these sites. Any patient who had an aortic measurement >4.5 cm with a sustained >1 cm reduction in their aortic measurements on follow-up scans were included in a separate intervention group. As an additional measure these individuals were cross referenced with free-text and common abbreviations in report indications and summaries for the terms “aortic valve / root replacement” and “post-operative”.

Using the International Classification of Disease 10^th^ Revision (ICD-10) the following causes of death were examined; ‘cardiovascular disease’(CVD) for any individual whose death was determined within ICD-10 chapters I00-I99 i.e. diseases of the circulatory system, ‘aortic death’ as any individual whose primary cause of death fell under the I71 code i.e. aortic aneurysm and dissection, and ‘thoracic aortic death’ for those whose primary cause of death fell in ICD-10 sub-codes I71.01, I71.1 or I71.2 i.e. aneurysm, dissection or rupture of the thoracic aorta.

### Statistical analysis & ethics

Statistical tests were performed to compare distributions between ECHO parameters, demographics, mortality and the intervention group. Continuous variables were assessed for normality of distribution and unpaired Wilcoxon rank sum tests were applied along with logistic regression analyses. For categorical variables, Pearson’s Chi-squared tests were used. The survival analyses were performed up to 10-year follow-up with different outcomes and group comparisons. Log rank tests were applied to compare differences between survival curves. Multivariate Cox proportional hazard regression models were used to evaluate the effect of variables on the survival functions. Adjustments included sex, age, weight, the presence of left ventricular hypertrophy, ejection fraction, aortic repair, and the presence of atrial fibrillation. The proportional hazards assumptions for all covariates were tested based on the scaled Schoenfeld residuals. For all statistical tests, p-value <0.01 was determined as significantly different. All data cleaning, processing, and statistical analyses were performed using R Statistical Software (v4.3.2, R Core Team, 2023).

NEDA is registered with the publicly accessible Australian New Zealand Clinical Trials Registry (ACTRN12617001387314). Ethical approval has been obtained from all relevant Human Research Ethics Committees (HREC) in each State and Territory of Australia. Datasets can be made available upon request to the corresponding author for the validation of the statistical methods and analyses.

## Results

### Descriptive, demographic & follow-up data

Demographic and echocardiographic data are presented in **Tables 1** and **2**. The mean age at baseline ECHO was 63±16 years, with a 56% male predominance. On average patients had undergone 3±2 studies with a mean follow-up time 7.3±4.4 years. The average size of the ascending aorta at any point was 3.4±0.5 cm, with females having smaller aortic dimensions at all ascending aortic sites (p<0.001, see **Table S1**). The prevalence of any thoracic aortic dilatation (>4 cm) across the cohort was 12% and 0.3% for those with severe aortic dilatation (>5 cm). SoV was the most frequently recorded ascending aortic measurement (476,541 measurements representing >99% of all scans) with a significant reduction in recorded values seen when moving from proximal to distal ascending aortic sites (STJ; 83,004 measurements, 17% of all scans and AscAo; 44,504 measurements, 9% of all scans; p<0.0001).

**Table 1.** Demographic and study characteristics separated by mortality.

| Characteristics | All<br>N = 175,158 | Outcome |  | p-value |
| --- | --- | --- | --- | --- |
|  |  | Alive<br>N = 119,297 | Dead<br>N = 55,861 |  |
| Sex |  |  |  |  |
| Male | 97,688 (56%) | 65,293 (55%) | 32,395 (58%) | <0.001<sup>1</sup> |
| Female | 77,470 (44%) | 54,004 (45%) | 23,466 (42%) | <0.001<sup>1</sup> |
| Age |  |  |  |  |
| At baseline scan, y | 62.7±16.3 | 58.6±16.2 | 71.3±13.0 | <0.001<sup>2</sup> |
| At last scan, y | 65.6±16.4 | 61.7±16.3 | 74.1±13.1 | <0.001<sup>2</sup> |
| Scans |  |  |  |  |
| Number of scans per patient | 2.7±2.0 | 2.7±2.0 | 2.7±1.9 | 0.3<sup>2</sup> |
| Follow up |  |  |  |  |
| Interval between scans, y | 2.0±2.0 | 2.1±2.1 | 1.8±1.9 | <0.001<sup>2</sup> |
| Interval from first to last scan, y | 3.0±3.3 | 3.1±3.4 | 2.8±3.1 | <0.001<sup>2</sup> |
| Total follow up time,* y | 7.3±4.3 | 8.2±4.2 | 5.3±3.9 | <0.001<sup>3</sup> |
| Intervention |  |  |  |  |
| Aortic repair | 1,215 (1%) | 839 (1%) | 376 (1%) | 0.5<sup>2</sup> |

<sup>1</sup> Pearson's Chi-squared test
<sup>2</sup> Wilcoxon rank sum test
y; years
\*; years from first scan to death/last NDI census date

**Table 2.**
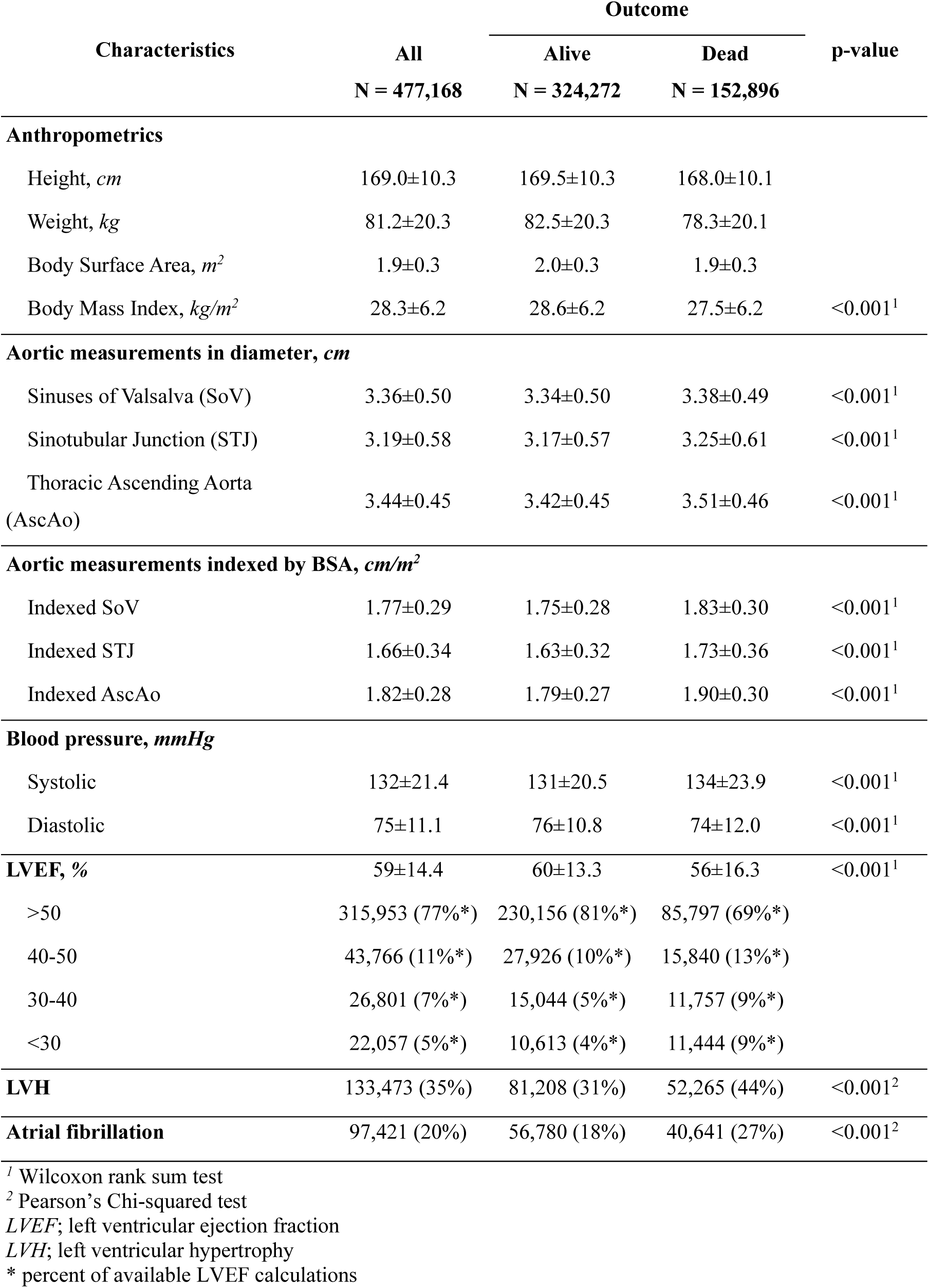
Overall echocardiographic and patient characteristics separated by mortality.

| Characteristics | All<br>N = 477,168 | Outcome |  | p-value |
| --- | --- | --- | --- | --- |
|  |  | Alive<br>N = 324,272 | Dead<br>N = 152,896 |  |
| Anthropometrics |  |  |  |  |
| Height, *cm* | 169.0±10.3 | 169.5±10.3 | 168.0±10.1 |  |
| Weight, *kg* | 81.2±20.3 | 82.5±20.3 | 78.3±20.1 |  |
| Body Surface Area, *m*<sup>2</sup> | 1.9±0.3 | 2.0±0.3 | 1.9±0.3 |  |
| Body Mass Index, *kg/m*<sup>2</sup> | 28.3±6.2 | 28.6±6.2 | 27.5±6.2 | <0.001<sup>1</sup> |
| Aortic measurements in diameter, *cm* |  |  |  |  |
| Sinuses of Valsalva (SoV) | 3.36±0.50 | 3.34±0.50 | 3.38±0.49 | <0.001<sup>1</sup> |
| Sinotubular Junction (STJ) | 3.19±0.58 | 3.17±0.57 | 3.25±0.61 | <0.001<sup>1</sup> |
| Thoracic Ascending Aorta<br>(AscAo) | 3.44±0.45 | 3.42±0.45 | 3.51±0.46 | <0.001<sup>1</sup> |
| Aortic measurements indexed by BSA, *cm/m*<sup>2</sup> |  |  |  |  |
| Indexed SoV | 1.77±0.29 | 1.75±0.28 | 1.83±0.30 | <0.001<sup>1</sup> |
| Indexed STJ | 1.66±0.34 | 1.63±0.32 | 1.73±0.36 | <0.001<sup>1</sup> |
| Indexed AscAo | 1.82±0.28 | 1.79±0.27 | 1.90±0.30 | <0.001<sup>1</sup> |
| Blood pressure, *mmHg* |  |  |  |  |
| Systolic | 132±21.4 | 131±20.5 | 134±23.9 | <0.001<sup>1</sup> |
| Diastolic | 75±11.1 | 76±10.8 | 74±12.0 | <0.001<sup>1</sup> |
| LVEF, % | 59±14.4 | 60±13.3 | 56±16.3 | <0.001<sup>1</sup> |
| >50 | 315,953 (77%*) | 230,156 (81%*) | 85,797 (69%*) |  |
| 40-50 | 43,766 (11%*) | 27,926 (10%*) | 15,840 (13%*) |  |
| 30-40 | 26,801 (7%*) | 15,044 (5%*) | 11,757 (9%*) |  |
| <30 | 22,057 (5%*) | 10,613 (4%*) | 11,444 (9%*) |  |
| LVH | 133,473 (35%) | 81,208 (31%) | 52,265 (44%) | <0.001<sup>2</sup> |
| Atrial fibrillation | 97,421 (20%) | 56,780 (18%) | 40,641 (27%) | <0.001<sup>2</sup> |

<sup>1</sup> Wilcoxon rank sum test<sup>2</sup> Pearson's Chi-squared test
LVEF; left ventricular ejection fraction
LVH; left ventricular hypertrophy
\* percent of available LVEF calculations

At the time of last census to NDI mortality data, 55,861 (32%) patients had died of any cause, with CVD mortality the most commonly reported cause (n=19,208, 34% of those who had died); more than double that of the second most common cause which was neoplasms (**Figure S1**). Aortic death was listed as the primary cause of death in 715 (1.3% of all deaths) and thoracic aortic death in 114 (0.2%). The incidence of aortic dilatation (>4 cm) was 10 cases per 1,000 person-years and 1 per 1,000 person-years for severe aortic dilatation. The incidence of any aortic death was 6 cases per 10,000 person-years and thoracic aortic death 1 per 10,000 person-years.

### All-cause mortality by aortic dimensions

All-cause mortality survival probabilities by severity of aortic dilatation at any site is shown in **Figure 2** and separated by specified aortic sites in **Figure S2**. Cox regression analyses for all-cause mortality by severity and site of thoracic dilatation is shown in **Table S2**. 10-year all-cause mortality was higher for those with severe aortic dilatation at any site compared to individuals with normal aortic diameters (52 vs. 36%, p<0.0001). Following age, sex and weight-adjusted multivariate regression analyses the likelihood of 10-year all-cause mortality with severe aortic dilatation at any site was increased compared to those with normal dimensions (HR 1.59, CI 1.22-2.07; p<0.001) and driven by the effect of CVD mortality on the population, which accounted for 49% of all deaths in those with severe TAA.

**Figure 2:**
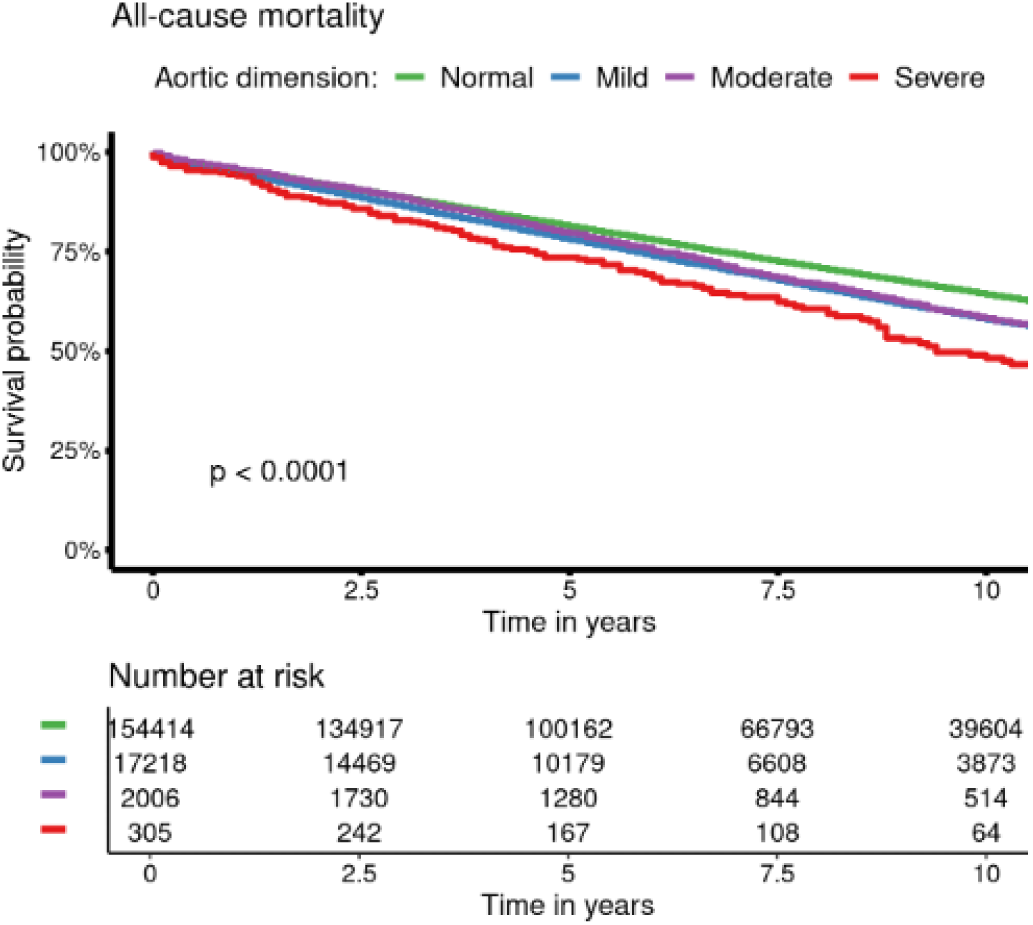
Survival probability for all-cause mortality by degree of aortic dilatation. Kaplan-Meier plot for all-cause mortality survival grouped by maximal aortic diameter from any ascending aortic site. There is a statistically significant reduction in survival probabilities for those with severe aortic dilatation compared to those with normal aortic dimensions (52 vs 36% mortality; p<0.0001).

### CVD mortality by aortic dimensions

Kaplan-Meier and multivariate Cox analysis for cardiovascular death by severity of aortic size and site are shown in **Figure 3** and **Table 3**; and as continuous variables in **Figure 4**. 10-year CVD mortality was greater with severe aortic dilatation at any site compared to those with normal aortic dimensions (31 vs. 14%, p<0.0001) (**Figure 3A**). When moving from the proximal to distal ascending aorta survival probability from CVD mortality worsened, SoV (30%; HR 1.79; CI 1.2-2.67; p=0.004), STJ (41%; HR 1.91; CI 1.11-3.29; p=0.002) and AscAo (45%; HR 3.96; CI 2.06-7.64; p<0.001) respectively (**Figure 3B-D** and **Table 3**). Increasing aortic size - both absolute and BSA-adjusted - corresponded with an increased probability of CVD mortality, with the ascending aorta conferring the highest probability of CVD death compared with the SoV and STJ (**Figure 4**). Older age and a BMI <17kg/m^2^ were independent predictors of CVD mortality and female sex and BMI >26kg/m^2^ was protective (**Table 3**). Other independent ECHO predictors of CVD mortality are shown in **Table S3** and included the presence of left ventricular hypertrophy, atrial fibrillation and worsening left ventricular function. BSA-adjusted CVD mortality data are shown in **Figure S3** and separated by gender in **Figure S4**.

**Figure 3:**
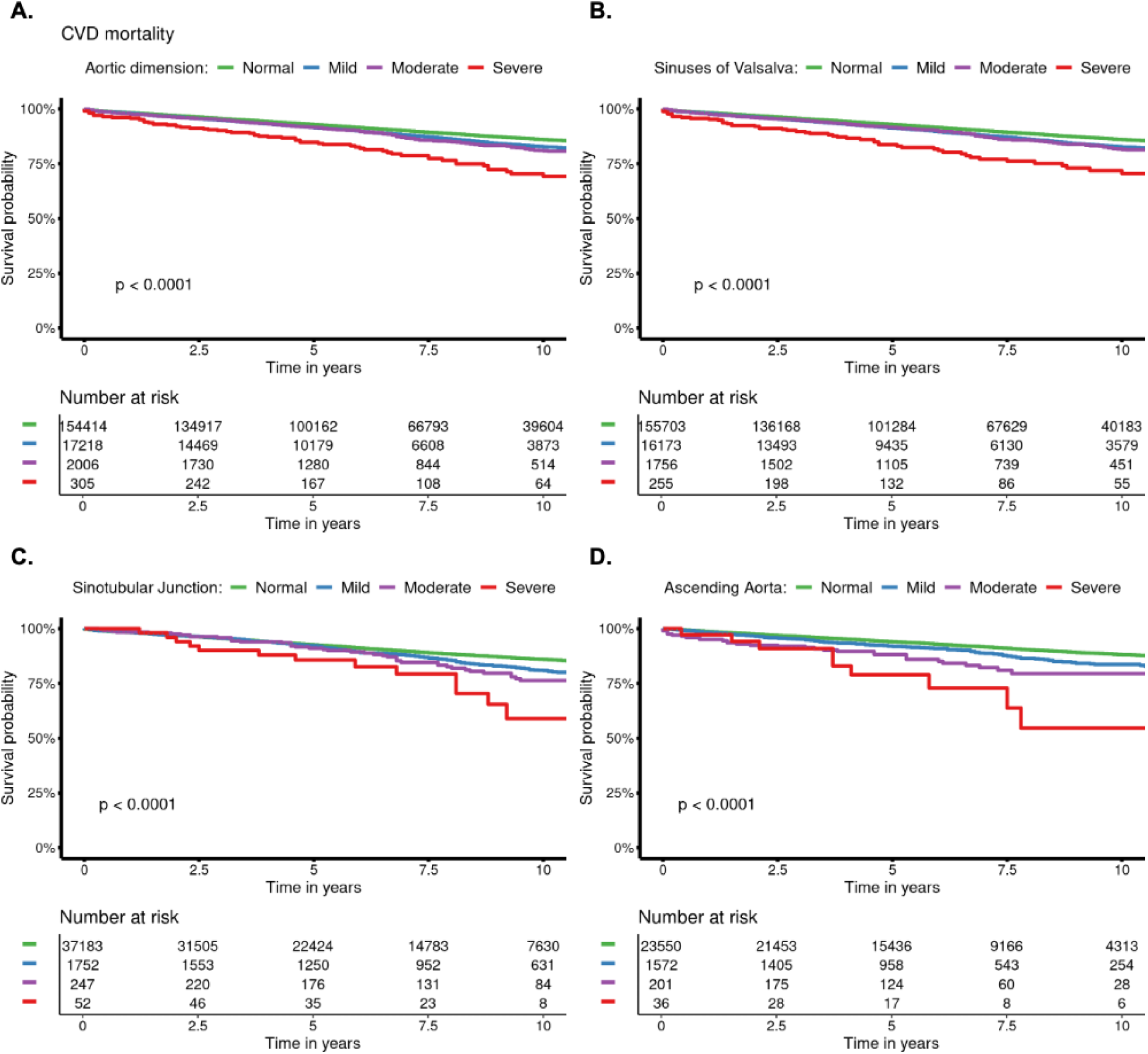
Survival probabilities for CVD mortality by degree of aortic dilatation at any site and separated by aortic site. Kaplan-Meier plots for CVD mortality grouped by (A) maximal aortic diameter at any site, and separated by specific ascending aortic sites (B-D). There is a statistically significant reduction in survival probabilities for those with severe dilatation compared to those with normal aortic dimensions at all aortic sites (30 vs. 17% mortality; p< 0.0001) with an incremental worsening of survival probabilities when moving from the proximal to distal thoracic ascending aorta. Severe aortic dilatation at the SoV (B) confers 30% 10-year CVD mortality risk compared to 41% at the STJ (C) and 45% at the AscAo (D).

**Table 3.**
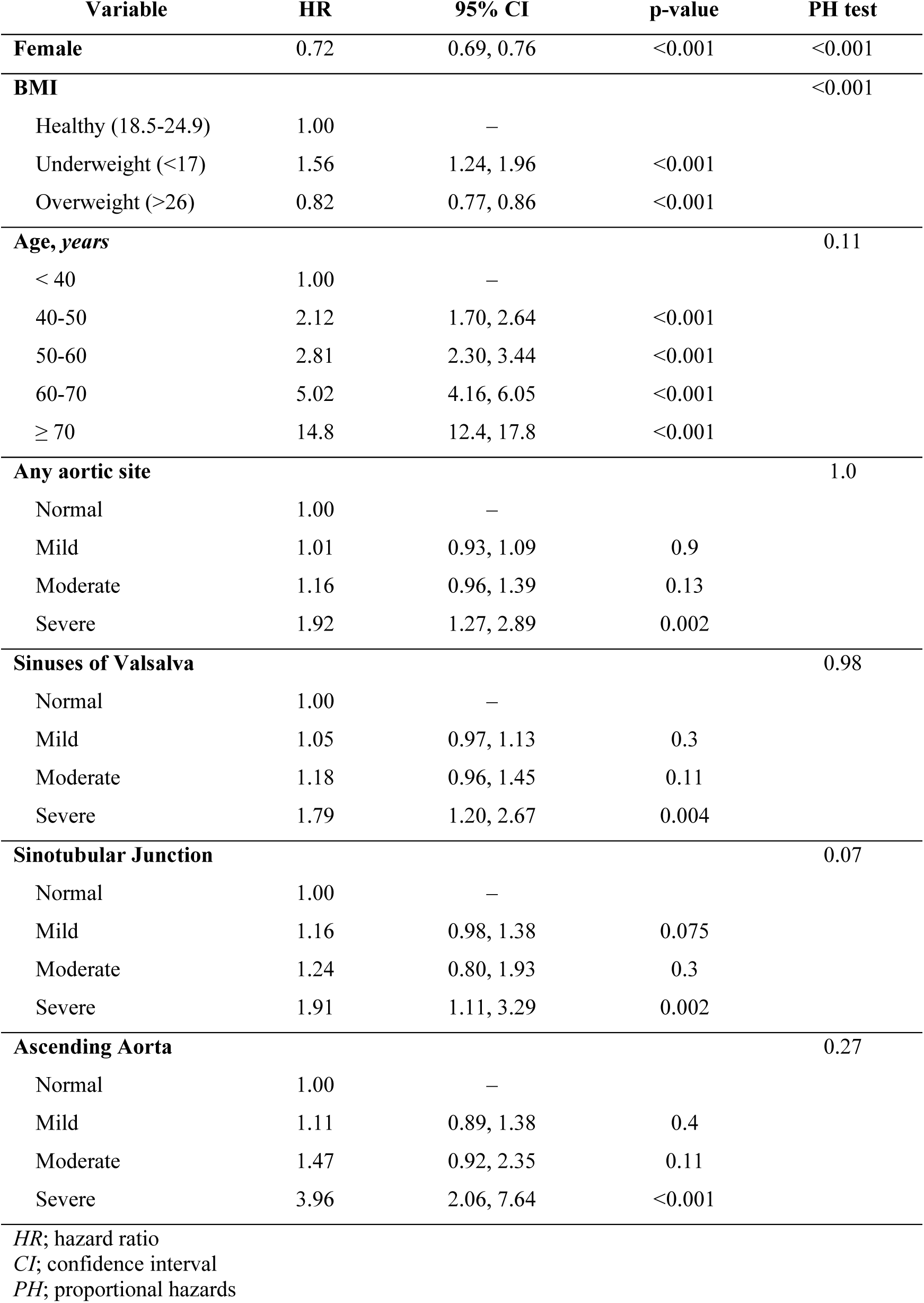
Cox regression analysis for cardiovascular mortality.

**Figure 4:**
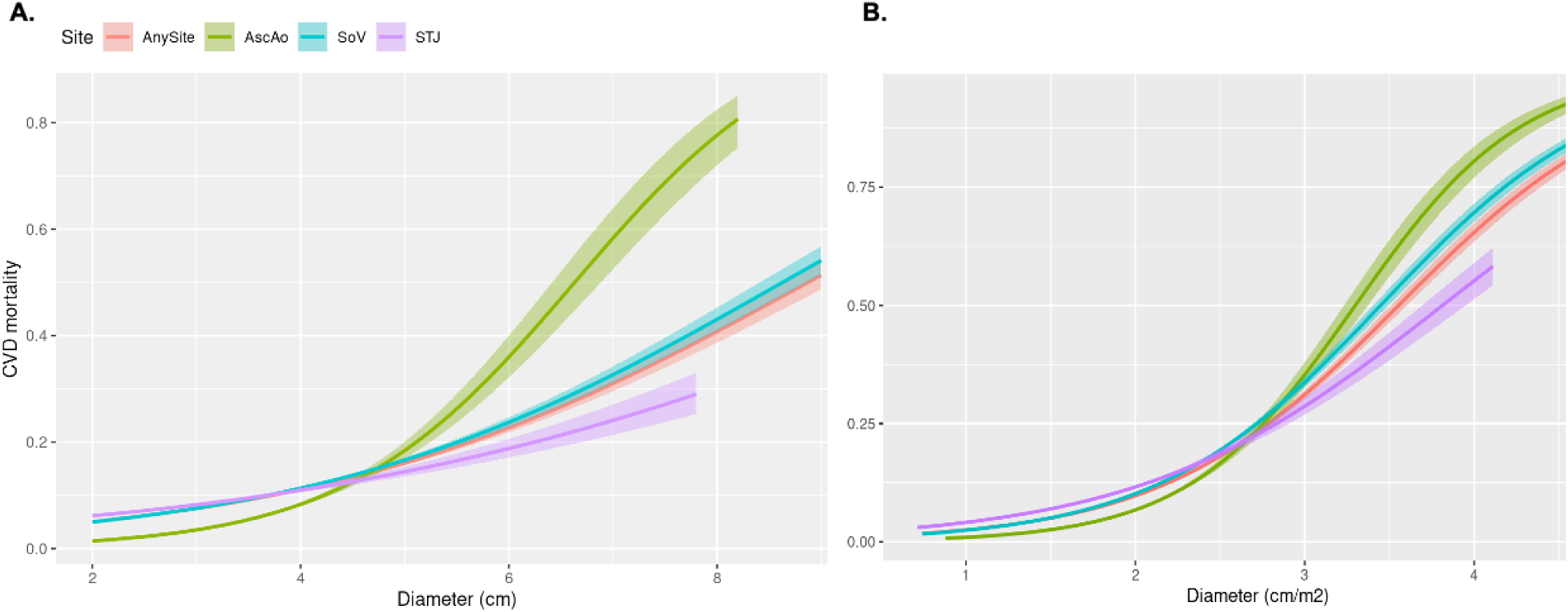
Logistic regression plots showing the probability of CVD mortality based on (A) Absolute and (B) BSA-adjusted aortic size separated by site. (A) there is progressive increase in the likelihood of CVD mortality with increasing absolute aortic size, and a notable inflection point for the AscAo beyond 5 cm when compared to the SoV and STJ. (B) Following BSA-adjustment there is partial normalisation of the respective curves, with a steeper increase in the probability of CVD mortality seen from 2.75 cm/m^2^ at all aortic sites.

### Aortic mortality by aortic dimensions

Despite an overall low incidence of aortic death across the cohort, severe TAA predicted aortic death at all ascending aortic sites compared to those with normal aortic diameters following multivariate analysis (**Table S4**). Similar to CVD mortality, other independent predictors of aortic death were a low BMI and older age. Severe TAA conferred a 54-fold greater risk of thoracic aortic death (OR 53.9, CI 24.39-119.43; p<0.0001) compared to those with normal aortic dimensions, despite the majority (>50%) of these deaths occurring in those with normal aortic diameters (**Figure 5**); in keeping with the aortic paradox and owing to the vast majority of the study population (>88%) having normal aortic diameters compared to <1% having severe aortic dilatation.

**Figure 5:**
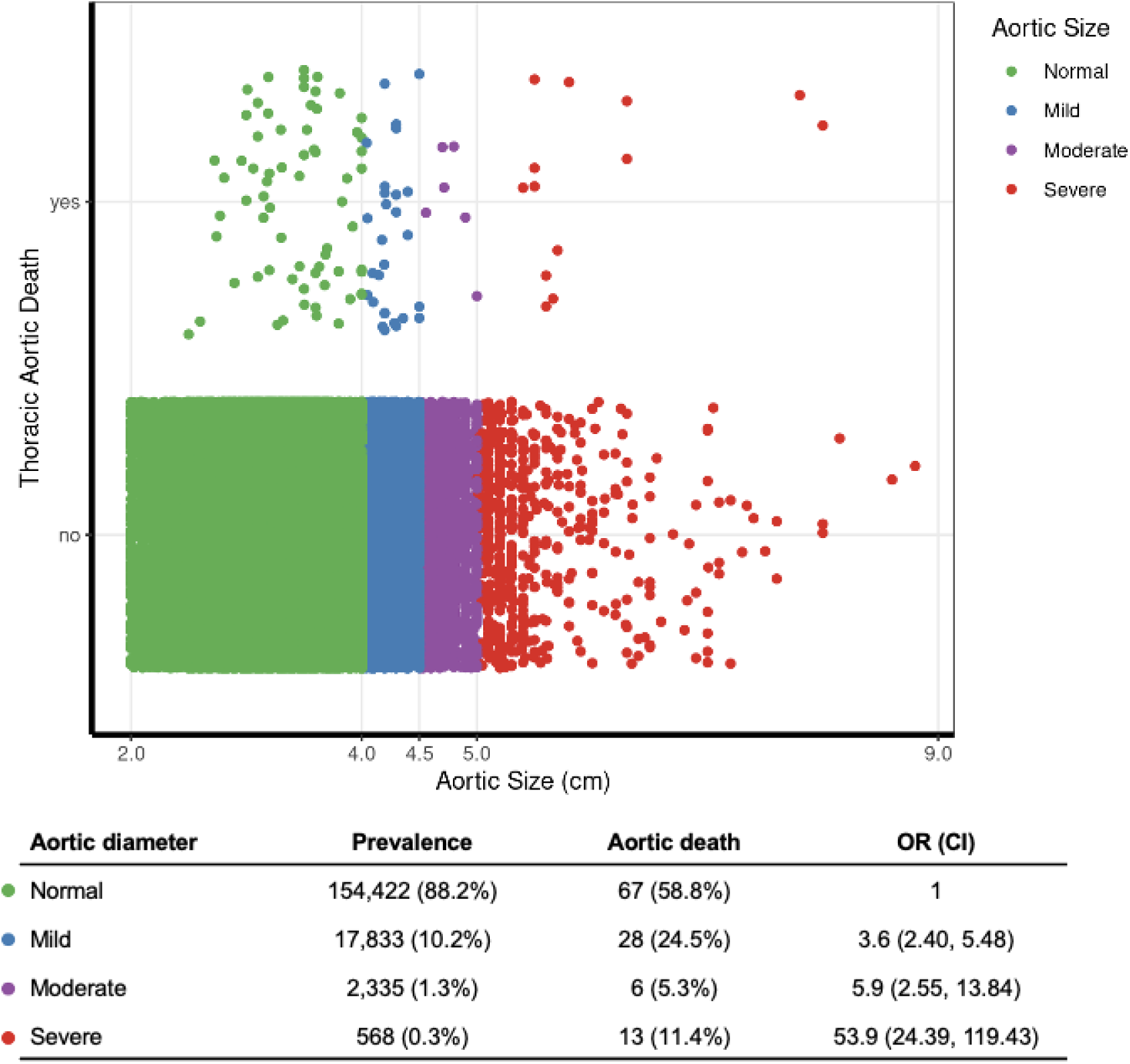
Scatter plot of thoracic aortic death by aortic size

### Effect of aortic intervention on mortality

Aortic repair balanced 10-year all-cause mortality probability to that seen in the population with normal aortic diameters (36 vs. 35%, p=0.2) (**Figure 6A**). Similarly, there was normalisation of 10-year CVD mortality (15 vs 14%, p=0.3) (**Figure 6B**), despite an increased risk of post-surgical aortic death following repair (HR 2.88; CI 1.62-5.11; p<0.001) (**Table S4**), which occurred in 5% of the intervention group compared to 0.5% of those with normal aortic dimensions who had not undergone aortic repair.

**Figure 6:**
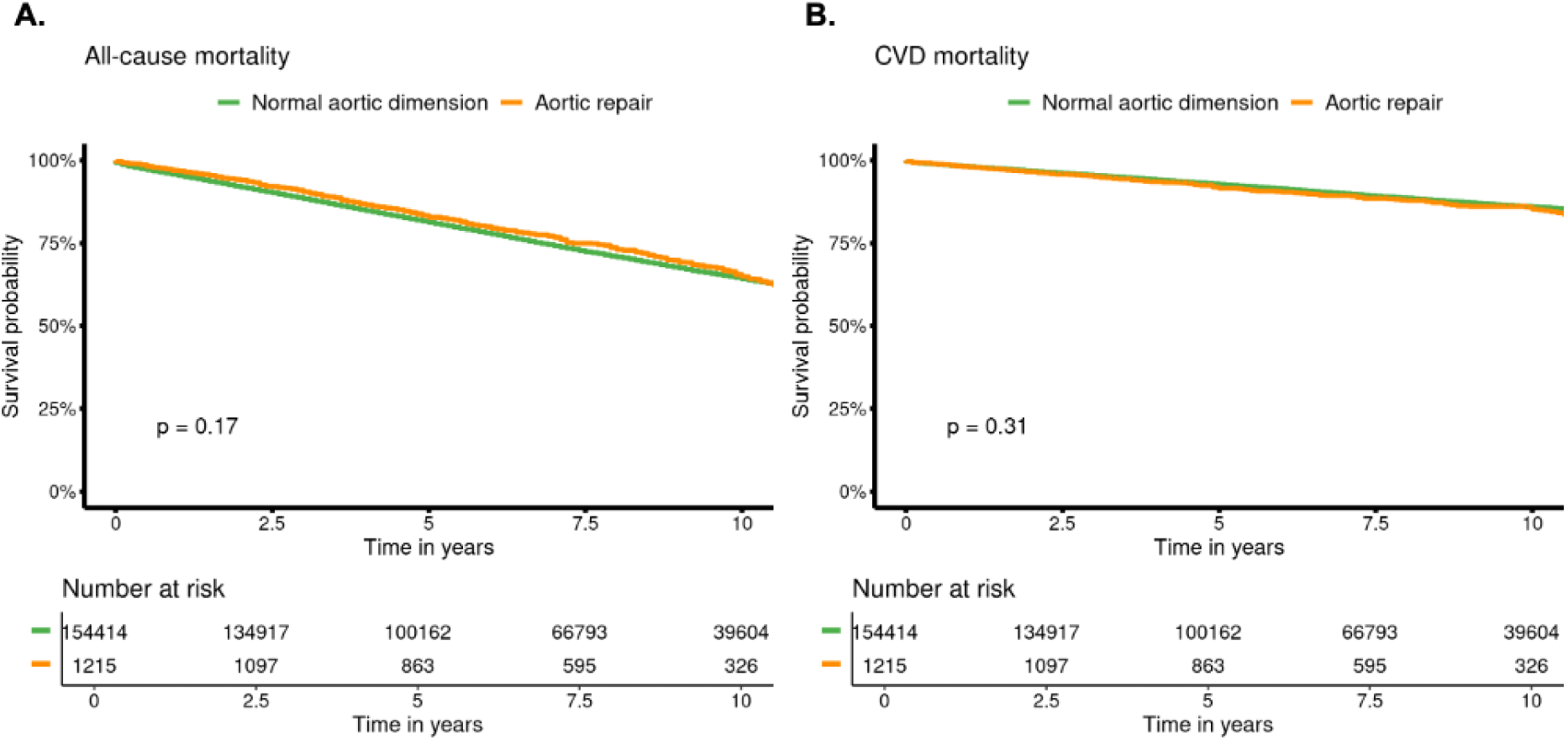
Survival probabilities for all-cause and CVD mortality following aortic intervention. Kaplan-Meier survival plots for (A) all-cause mortality and (B) CVD mortality comparing those having undergone surgical aortic intervention to those with normal aortic dimensions. Both 10-year all-cause (35 vs. 36%; p=0.17) and CVD mortality (15 vs. 14%; p=0.31) are comparable between those who underwent aortic repair and the population with healthy aortic diameters.

## Discussion

In this study which examined a nationwide echocardiographic database linked to population mortality records it has been shown that severe ascending aortic dilatation at any site confers a 2-fold increase in 10-year CVD mortality compared to those with normal aortic dimensions. These results are not explained by the effect of AoD on the cohort given the overall low event rate of thoracic aortic death in the study population which accounted for just 0.2% (1 per 10,000 patient-years) of all deaths.

In addition, the risk of CVD death with severe TAA increases when moving from the proximal to distal ascending aortic sites, with severe AscAo aneurysms doubling the likelihood of CVD mortality compared to severe aortic root aneurysms. These findings are novel with prognostic implications for patients with aortopathy and the clinicians who care for them, and are made more noteworthy by the finding that surgical aortic repair augments the risk of mortality to that of the healthy aortic population.

In the present study an aortic dimension >5 cm was selected as the cut-off for severe aortic dilatation, less than the prescribed 5.5 cm barrier to preventative surgical correction stated in some guidelines.(14, 15) Recent data has demonstrated that unoperated patients with aortic diameters 5-5.5 cm are at higher risk of adverse aortic events than previously suspected, with similar rates of adverse aortic events to those seen at diameters 5.5-6 cm.(26) Our results support these findings, identifying an ascending aortic measurements >5 cm (in particular at the ascending aorta) as conferring greater risk of mortality when compared to those with aortic measurements of <5 cm, and are in-line with some of the more contemporaneous guideline recommendations.(16, 17)

The prevalence of severe TAA and incidence of aortic death are at the higher range of what has been reported in population studies.(10–13) These results are likely owing to the older age and co-morbidities in this echocardiographic cohort. Patients referred for echocardiograms are more likely to have some cardiovascular disorder underlying their reason for referral, therefore it is reasonable to assume that the prevalence of CVD processes which drive aneurysm formation and progression are higher within NEDA than across the wider population.

The underlying reason for the association between aortic disease severity and CVD mortality is likely multi-factorial. Aortic aneurysms are a general indicator of poor cardiovascular status and co-morbidity and as such may represent a correlate of poorer CVD status. Given the older mean age of the cohort and the knowledge that aneurysms in older populations are driven by factors such as aging, hypertension, smoking and atherosclerotic disease burden,(27, 28) the presence of aortic dilatation may simply reflect the presence of concomitant traditional CVD risk factors. However, the appreciable worsening of CVD survival probability seen in those with severe compared to moderate dilatation suggests that severe aneurysms may play a more direct role in CVD outcomes.

It is probable that another factor driving the higher rates of CVD mortality seen in severe aortic disease is due to the under-recognition of AoD as the primary cause of death in some of these cases. The under reporting of AoD is well established and relates to a number of factors. Firstly, acute AoD is rapidly fatal with 40% dying immediately,(29, 30) and when death occurs prior to hospitalisation it may be erroneously attributed to more common cardiovascular causes. Secondly the most common symptom of AoD is chest pain,(31) which if reported pre-mortem may be attributed to acute myocardial infarction in the absence of post-mortem examination. Thirdly, AoD is frequently missed on initial presentation,(32, 33) and early mortality from its sequelae may be misclassified as non–dissection related. Lastly, given the older average age of AoD presentations (63 years) and male predominance,(34) coupled with the aforementioned commonly associated CVD co-morbidities, post-mortem examination may be less frequently pursued even when some degree of aortic dilatation is known ante-mortem.

The finding of worse CVD outcomes in those with severe ascending aortic dilatation compared to severe dilatation at the sinus of Valsalva and sinotubular junction is thought-provoking. This may be explained by the comparative smaller sample sizes at the STJ and AscAo, or due to the underlying processes and biomechanics driving aneurysm progression at these sites. AscAo aneurysms may be more susceptible to systemic factors driving progression such as hypertension, shear wall stress, aortic distensibility and overall plaque burden.(35–37) Additionally, AoD at the AscAo is more likely to involve the aortic arch and supra-aortic vessels with a resultant increased risk of neurological malperfusion and CVD mortality. This has been demonstrated in a German registry of type A dissection which found that 20% of patients presenting with AoD had neurological dysfunction that was ultimately associated with extensive malperfusion syndrome and mortality.(38) Importantly, current guidelines do not differentiate in their recommendations for surgical correction of proximal TAA depending on site.(14, 15) Our data supports further investigation into whether proximal thoracic aortic aneurysm site confers disparate prognostic risk, and a such should be considered differently.

The use of an intervention group to demonstrate a reduction in all-cause and CVD mortality to that of a population with normal aortic parameters is interesting, though warrants scrutiny. Firstly, confirmation of definite aortic repair could not be validated in each case, instead a surrogate of a significant and sustained reduction in absolute aortic size was correlated with free-text report indications and summaries commonly seen in those who have undergone ascending thoracic aortic repair to produce the intervention arm. Secondly, perioperative mortality in those failing to reach post-operative echocardiography cannot be accounted for by our analysis. Thirdly, it is likely that concomitant coronary artery bypass grafting and aortic valve replacement was performed in many cases of aortic intervention, and confounding from these procedures cannot be excluded as the reason for the balancing of CVD mortality in this study. Nevertheless, given that contemporary clinical work-up results in coronary revascularisation and/or valve replacement being performed in concert with aortic surgery these factors should not necessarily limit the real-world applicability of the results.

There are some additional limitations that need to considered. Due to the retrospective nature of the study and exclusive utilisation of echocardiographic data it was not possible to include all relevant variables. An additional limitation is the absence of outcome data beyond mortality. Our methodology of linkage with the National Death Index captured only episodes of fatal aortic dissection, and not those in which the aortic dissection was survived. The addition of morbidity, surgical, genetic, patient history and medication data would strengthen the depth of analysis, minimising confounding and improving the overall reliability of the results.

Irrespective of correlative or causative role underlying the present findings, the results of this study are novel and provide real-world pre-mortem data that can be applied for prognostication of patients with ascending aortic aneurysms; informing subsequent prospective studies, future guidelines and aiding in the personalisation of management beyond absolute aortic size. Continued exploration of imaging predictors of TAA and AoD are imperative to reduce adverse outcomes and guide the timing of surgical intervention.

In conclusion, through the analysis of over 175,000 patients and almost 500,000 echocardiograms linked to mortality data it has been demonstrated that severe ascending aortic disease portends a higher likelihood of 10-year CVD mortality and that the risk of this outcome incrementally increases when moving from the proximal to distal ascending aorta. In addition, aortic repair comparatively reduces the risk of mortality in those with aortic aneurysm, balancing survival rates to that seen in a healthy aortic population. Prospective observational studies in this high-risk groups exploring the differences in outcomes of patients depending on aneurysm location will aid in the further translation of these novel findings.

## Data Availability

Datasets can be made available upon request to the corresponding author for the validation of the statistical methods and analyses.

## Supplemental Figures

**Figure S1:**
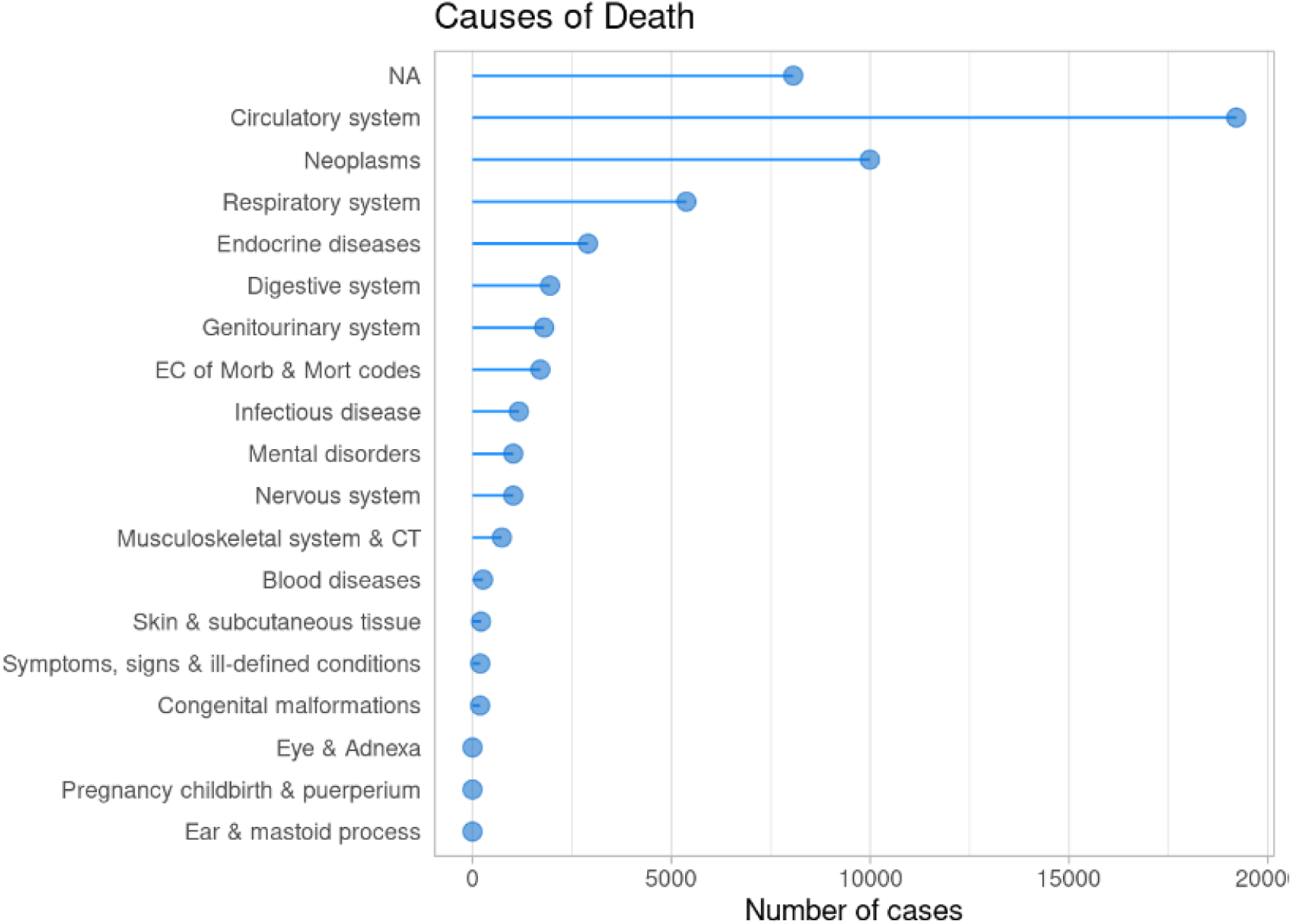
Distribution of causes of death by NDI chapter.

**Figure S2:**
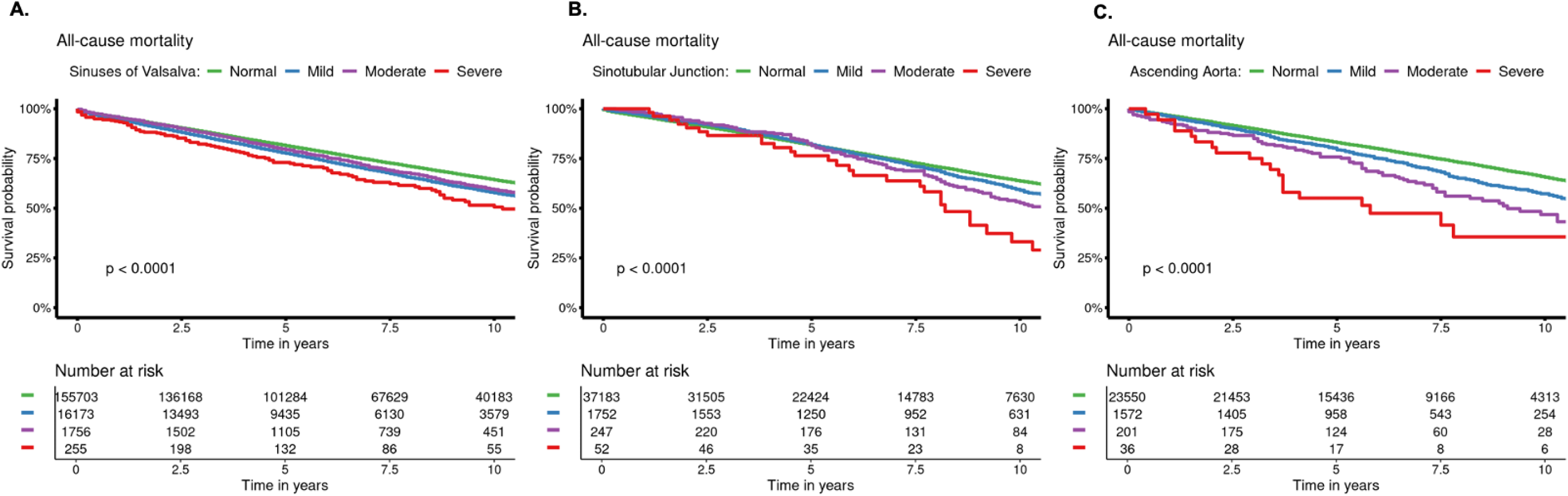
Survival probability for all-cause mortality by degree of aortic dilatation at specified aortic sites

**Figure S3:**
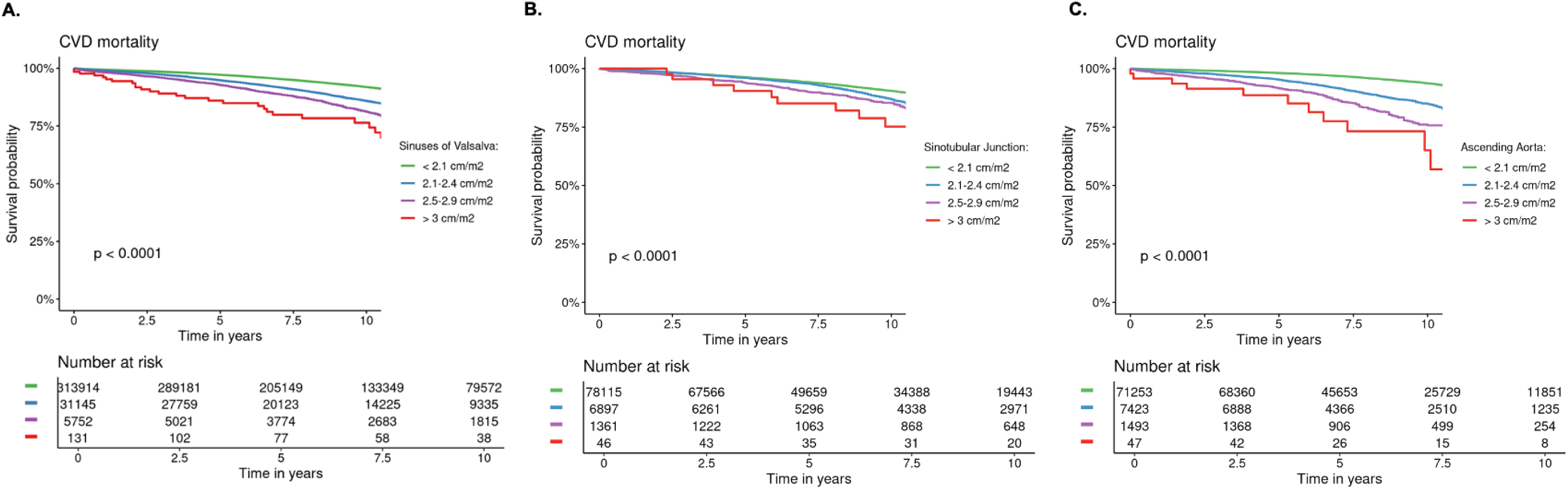
Survival probability for CVD mortality by BSA-adjusted aortic dimensions specified by aortic sites

**Figure S4:**
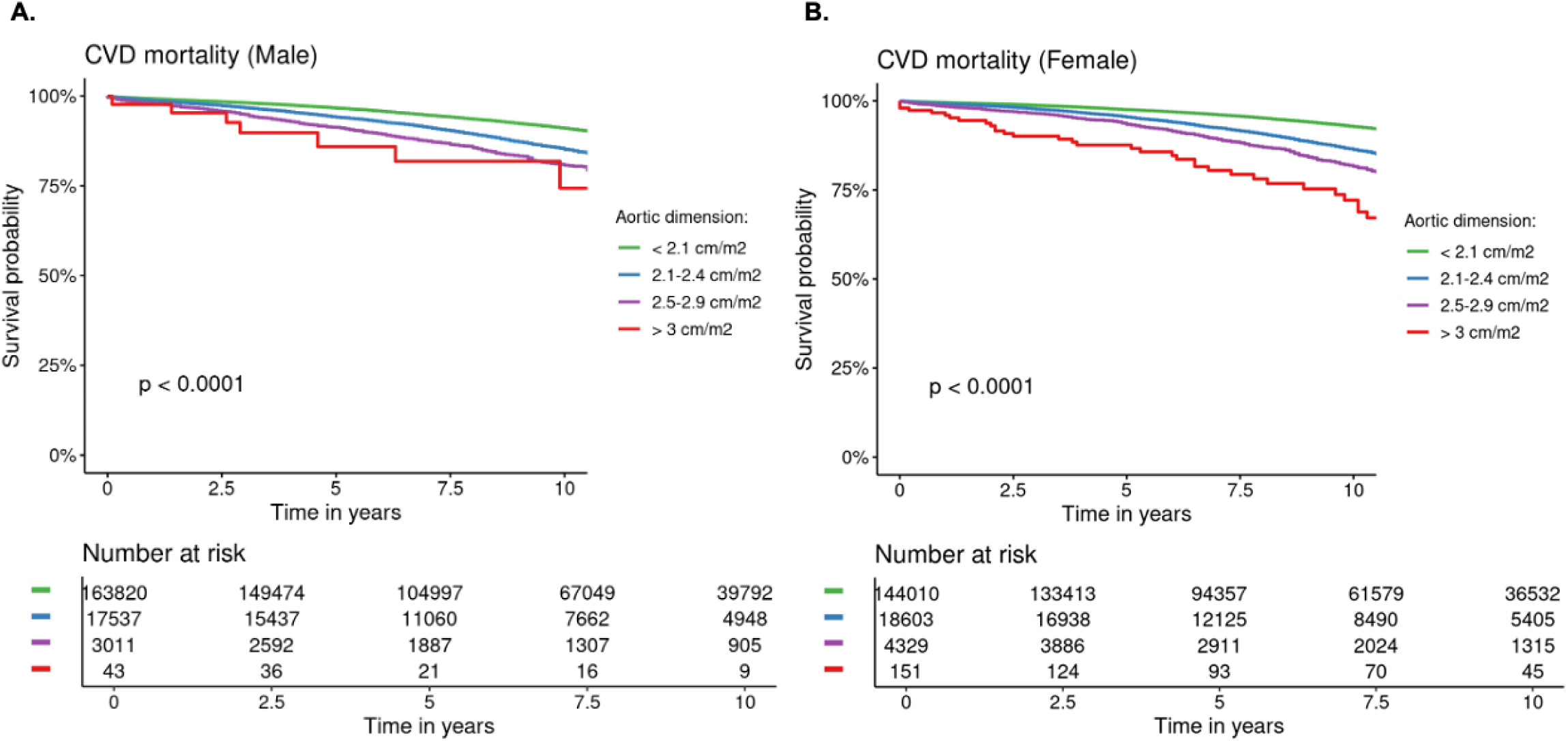
Survival probability for CVD mortality by BSA-adjusted aortic dimensions at any site separated by gender

## Supplemental Tables

**Table S1.** Mean absolute and BSA corrected aortic measurements at baseline.

| Measurement | Number | Female | Male | p-value<sup>/</sup> |
| --- | --- | --- | --- | --- |
| **Absolute diameter, *cm*** |  |  |  |  |
| SoV | 174,888 | 3.08±0.41 | 3.57±0.48 | <0.001 |
| STJ | 25,859 | 3.04±0.57 | 3.29±0.57 | <0.001 |
| AscAo | 17,410 | 3.27±0.43 | 3.56±0.44 | <0.001 |
| **Indexed to body surface area, *cm/m<sup>2</sup>*** |  |  |  |  |
| Indexed SoV | 108,869 | 1.76±0.30 | 1.78±0.28 | <0.001 |
| Indexed STJ | 25,228 | 1.72±0.36 | 1.62±0.31 | <0.001 |
| Indexed AscAo | 17,402 | 1.86±0.31 | 1.77±0.26 | <0.001 |

<sup>/</sup> Wilcoxon rank sum test

**Table S2.** Cox regression analysis for all-cause mortality.

| Variable | HR | 95% CI | p-value | PH test |
| --- | --- | --- | --- | --- |
| **Female** | 0.74 | 0.72, 0.77 | <0.001 | <0.001 |
| **BMI** |  |  |  | <0.001 |
| Healthy (18.5-24.9) | 1.00 | – |  |  |
| Underweight (<17) | 1.70 | 1.49, 1.93 | <0.001 |  |
| Overweight (>26) | 0.84 | 0.81, 0.87 | <0.001 |  |
| **Age, years** |  |  |  | <0.001 |
| < 40 | 1.00 | – |  |  |
| 40-50 | 1.83 | 1.63, 2.04 | <0.001 |  |
| 50-60 | 2.49 | 2.25, 2.75 | <0.001 |  |
| 60-70 | 4.16 | 3.79, 4.57 | <0.001 |  |
| ≥ 70 | 10.2 | 9.31, 11.1 | <0.001 |  |
| **Any aortic site** |  |  |  | 0.51 |
| Normal | 1.00 | – |  |  |
| Mild | 1.02 | 0.97, 1.07 | 0.4 |  |
| Moderate | 1.01 | 0.90, 1.14 | 0.8 |  |
| Severe | 1.59 | 1.22, 2.07 | <0.001 |  |
| **Sinuses of Valsalva** |  |  |  | 0.66 |
| Normal | 1.00 | – |  |  |
| Mild | 1.06 | 1.01, 1.11 | 0.013 |  |
| Moderate | 1.03 | 0.91, 1.17 | 0.6 |  |
| Severe | 1.75 | 1.27, 2.39 | <0.001 |  |
| **Sinotubular Junction** |  |  |  | 0.01 |
| Normal | 1.00 | – |  |  |
| Mild | 0.95 | 0.86, 1.06 | 0.4 |  |
| Moderate | 1.08 | 0.82, 1.42 | 0.6 |  |
| Severe | 1.34 | 0.76, 2.36 | 0.3 |  |
| **Ascending Aorta** |  |  |  | 0.25 |
| Normal | 1.00 | – |  |  |
| Mild | 1.03 | 0.91, 1.17 | 0.7 |  |
| Moderate | 1.20 | 0.90, 1.61 | 0.2 |  |
| Severe | 2.25 | 1.38, 3.68 | 0.001 |  |

*HR*; hazard ratio*CI*; confidence interval*PH*; proportional hazards

**Table S3.** Cox regression analysis of other ECHO predictors of cardiovascular mortality.

| **Variable** | **HR** | **95% CI** | **p-value** | **PH test** |
| --- | --- | --- | --- | --- |
| **Aortic repair** | 1.28 | 0.89, 1.83 | 0.2 | 0.54 |
| **LVH** | 1.78 | 1.65, 1.92 | <0.001 | 0.23 |
| **LVEF, %** |  |  |  | 0.03 |
| > 50 | 1.00 | — |  |  |
| 40-50 | 1.62 | 1.49, 1.75 | <0.001 |  |
| 30-40 | 2.21 | 2.06, 2.37 | <0.001 |  |
| < 30 | 3.73 | 3.49, 3.98 | <0.001 |  |
| **Atrial fibrillation** | 1.35 | 1.24, 1.46 | <0.001 | 0.08 |

*HR*; hazard ratio*CI*; confidence interval*PH*; proportional hazards*LVEF*; left ventricular ejection fraction*LVH*; left ventricular hypertrophy

**Table S4.** Cox regression analysis for aortic death

| Variable | HR | 95% CI | p-value | PH test |
| --- | --- | --- | --- | --- |
| **Aortic repair** | 2.88 | 1.62, 5.11 | <0.001 | 1.00 |
| **Female** | 0.82 | 0.69, 0.98 | 0.031 | 0.96 |
| **BMI** |  |  |  | 0.99 |
| Healthy (18.5-24.9) | 1.00 | — |  |  |
| Underweight (<17) | 2.91 | 1.07, 7.93 | 0.037 |  |
| Overweight (>26) | 0.75 | 0.55, 1.02 | 0.068 |  |
| **Age, years** |  |  |  | 0.77 |
| < 40 | 1.00 | — |  |  |
| 40-50 | 1.22 | 0.50, 3.00 | 0.7 |  |
| 50-60 | 1.50 | 0.69, 3.28 | 0.3 |  |
| 60-70 | 3.94 | 1.98, 7.87 | <0.001 |  |
| ≥ 70 | 15.6 | 8.06, 30.3 | <0.001 |  |
| **Any aortic site** |  |  |  | 0.13 |
| Normal | 1.00 | — |  |  |
| Mild | 1.16 | 0.85, 1.59 | 0.4 |  |
| Moderate | 1.73 | 0.92, 3.26 | 0.088 |  |
| Severe | 5.96 | 2.22, 16.0 | <0.001 |  |
| **Sinuses of Valsalva** |  |  |  | 0.38 |
| Normal | 1.00 | — |  |  |
| Mild | 1.29 | 0.94, 1.77 | 0.12 |  |
| Moderate | 1.88 | 0.97, 3.66 | 0.063 |  |
| Severe | 4.32 | 1.07, 17.4 | 0.039 |  |
| **Sinotubular Junction** |  |  |  | 0.51 |
| Normal | 1.00 | — |  |  |
| Mild | 0.71 | 0.29, 1.75 | 0.5 |  |
| Moderate | 2.53 | 0.80, 8.01 | 0.11 |  |
| Severe | 5.30 | 5.63, 41.5 | <0.001 |  |
| **Ascending Aorta** |  |  |  | 0.66 |
| Normal | 1.00 | — |  |  |
| Mild | 1.13 | 0.49, 2.61 | 0.8 |  |
| Moderate | 2.99 | 0.73, 12.2 | 0.13 |  |
| Severe | 9.72 | 1.34, 70.3 | 0.024 |  |

*HR*; hazard ratio*CI*; confidence interval*PH*; proportional hazards

